# An exploratory assessment of the impact of a novel risk assessment test on breast cancer clinic waiting times and workflow: a discrete event simulation model

**DOI:** 10.1101/2022.06.13.22276333

**Authors:** Alison F. Smith, Samuel N. Frempong, Nisha Sharma, Richard D. Neal, Louise Hick, Bethany Shinkins

## Abstract

**Background:** Breast cancer clinics across the UK have long been struggling to cope with high demand. Novel risk prediction tools – such as the PinPoint test – could help to reduce unnecessary clinic referrals. Using early data on the expected accuracy of the test, we explore the potential impact of PinPoint on: (a) the percentage of patients meeting the two-week referral target, and (b) the number of clinic ‘overspill’ appointments generated.

**Methods:** A simulation model was built to reflect the annual flow of patients through a single UK clinic. Due to current uncertainty around the exact impact of PinPoint testing on standard care, two primary scenarios were assessed. Scenario 1 assumed complete GP adherence to testing, with only non-referred cancerous cases returning for delayed referral. Scenario 2 assumed GPs would overrule 20% of low-risk results, and that 10% of non-referred non-cancerous cases would also return for delayed referral. A range of sensitivity analyses were conducted to explore the impact of key uncertainties on the model results. Service reconfiguration scenarios, removing individual weekly clinics from the clinic schedule, were also explored.

**Results:** Under standard care, 66.3% (95% CI: 66.0 to 66.5) of patients met the referral target, with 1,685 (1,648 to 1,722) overspill appointments. Under both PinPoint scenarios, >98% of patients met the referral target, with overspill appointments reduced to between 727 (707 to 746) [Scenario 1] and 886 (861 to 911) [Scenario 2]. The reduced clinic demand was sufficient to allow removal of one weekly low-capacity clinic [N=10], and the results were robust to sensitivity analyses.

**Conclusions:** The findings from this early analysis indicate that risk prediction tools could have the potential to alleviate pressure on cancer clinics, and are expected to have increased utility in the wake of heightened pressures resulting from the COVID-19 pandemic. Further research is required to validate these findings with real world evidence; evaluate the broader clinical and economic impact of the test; and to determine outcomes and risks for patients deemed to be low-risk on the PinPoint test and therefore not initially referred.

## Background

The “two-week wait” (TWW) pathway for breast cancer, which stipulates that patients with suspected cancer symptoms should be seen in secondary care within two weeks of their first GP presentation, was first introduced in the UK in 1999 in response to the country’s poor breast cancer mortality statistics (1). Based on concerns over the number of cancer cases continuing to be identified via routine “symptomatic” referrals (i.e. where breast cancer is not initially suspected) (2), the TWW pathway was subsequently extended to all patients with breast symptoms (i.e. not limited to recognised cancer symptoms) (3). Similar TWW referral pathways are now in place for all major cancers, with over two million TWW referrals occurring annually across England alone (4).

The last five years has seen a sharp rise in TWW referrals for breast cancer, from just under 542,000 across England in 2015/16, to over 612,000 in 2019/20 (a rise of almost 13%) (5, 6). Unsurprisingly, breast cancer clinics are struggling to cope with the increased demand. Since 2018, the government’s operational standard target – that 93% of patients should be seen within fourteen days of referral – has not been achieved, and the ongoing COVID-19 pandemic has resulted in a further drop in recent adherence to this target (6-9). At the same time, whilst regional variation in cancer prevalence rates exists, less than 7% of patients referred along TWW cancer referral pathways are ultimately diagnosed with cancer, suggesting that many of these referrals could be avoided (10). Strategies to identify patients who do not require further investigation are urgently required to alleviate pressure on breast cancer clinics.

In collaboration with the Leeds Teaching Hospitals NHS Trust (LTHT) and the University of Leeds, PinPoint Data Science Ltd have developed a risk assessment tool, which is designed to determine patients’ risk of breast cancer based on a number of routine blood tests (including haematological, biochemical and tumour markers). These individual tests are combined within an algorithm (henceforth referred to as the “PinPoint test”) to provide a calibrated risk probability of cancer (a score between zero and one, with higher values indicating higher risk) (11). The PinPoint test can be undertaken around the time of referral, providing utility for two main use-cases: (1) as a rule-out test for patients with very low risk of cancer (i.e. avoiding secondary care referrals in this group); and (2) as a tool to prioritise high risk patients (i.e. fast-tracking referrals for these patients). Thus far, early diagnostic accuracy evidence suggests that for use-case (1) the test could avoid 20% of unnecessary referrals, thus freeing up secondary care resources to focus on expediated diagnosis for those most at risk (12).

Prior to commencement of the current study, initial discussions were undertaken with local commissioners at Leeds (the Leeds Clinical Commissioning Group [CCG]), in order to determine decision makers’ primary requirements for adopting new interventions in this field. Based on that work, the ability of new technologies to alleviate increasing pressure on secondary care services was identified as a primary concern for clinical decision makers and commissioners. The aim of this early exploratory evaluation was therefore to assess the potential impact of the PinPoint test on the flow of patients through clinic services (focusing on use-case (1)), evaluating two primary outcomes: (i) the percentage of patients seen in the clinic in under two weeks, and (ii) the number of ‘overspill’ appointments generated (i.e. where patients have to return to the clinic for further diagnostics due to insufficient same-day clinic capacity). Together these outcomes represent how well the referral system achieves timely diagnoses for patients within an efficiently functioning system.

Discrete event simulation (DES) is a useful modelling technique which enables simulation of individual entities (e.g. patients) through complex systems of activities (e.g. hospital services). With the inclusion of activities dependent on constrained resources (e.g. staff, rooms, devices), the possibility of queues forming in the system, leading to patient delays, can also be captured. Whilst predominantly used in the context of manufacturing and engineering, the use of DES in healthcare has been rising – with common applications including systems operation research, and disease progression modelling (13, 14). In the context of breast cancer services, most DES applications to date have focused on identifying optimal timings and/or technologies for breast cancer screening programs, without consideration of capacity constraints (15-19). Notable exceptions include two evaluations of mammography facilities – one in Brazil (20), the other USA (21) – which each modelled the flow of patients through mammography services to determine optimal routine staffing and equipment compositions. Similar methods are applied herein to instead explore the potential utility of a new intervention, the PinPoint test, for improving patient workflow in secondary care breast cancer clinics.

## Methods

Our reporting of the study methods below adheres to good practice guidelines as outlined in the Strengthening The Reporting of Empirical Simulation Studies (STRESS) checklist for DES models (22). Please see **Additional File 4** for the completed STRESS-DES checklist.

### Model Structure

A DES model was constructed in Simul8 (https://www.simul8.com) to reflect the flow of patients through the Leeds Teaching Hospitals Trust (LTHT) breast cancer clinic: a medium-to-large sized clinic which sees around 10,500 patients along the TWW pathway annually. The model tracks individual patients from their initial GP presentation, through to clinic services (including initial assessment, mammogram, ultrasound and biopsy), accounting for available clinic resources (e.g. staff and clinic rooms). Patients exit the model with a final diagnosis of breast cancer (following a multi-disciplinary team [MDT] meeting) or no breast cancer.

In the standard care arm (see **Figure 1** for the model schematic), all patients receive a referral to the clinic at their initial GP appointment; whilst in the PinPoint arm, referrals are based on whether patients receive a low- or high-risk PinPoint test result (see **Figure 2** for an illustration of the implementation pathway for the PinPoint test). Due to current uncertainty around the exact impact of PinPoint testing on standard care (particularly around the management and behaviour of patients with low-risk results), different scenarios are explored (see the *Analysis* section). Depending on the number of patients already in the queue for the clinic, referred patients have to wait for a period of time for the next available slot; similarly once in the clinic, patients must wait for the required staff and rooms to be available in order to undergo their required investigations.

**Figure 1.**
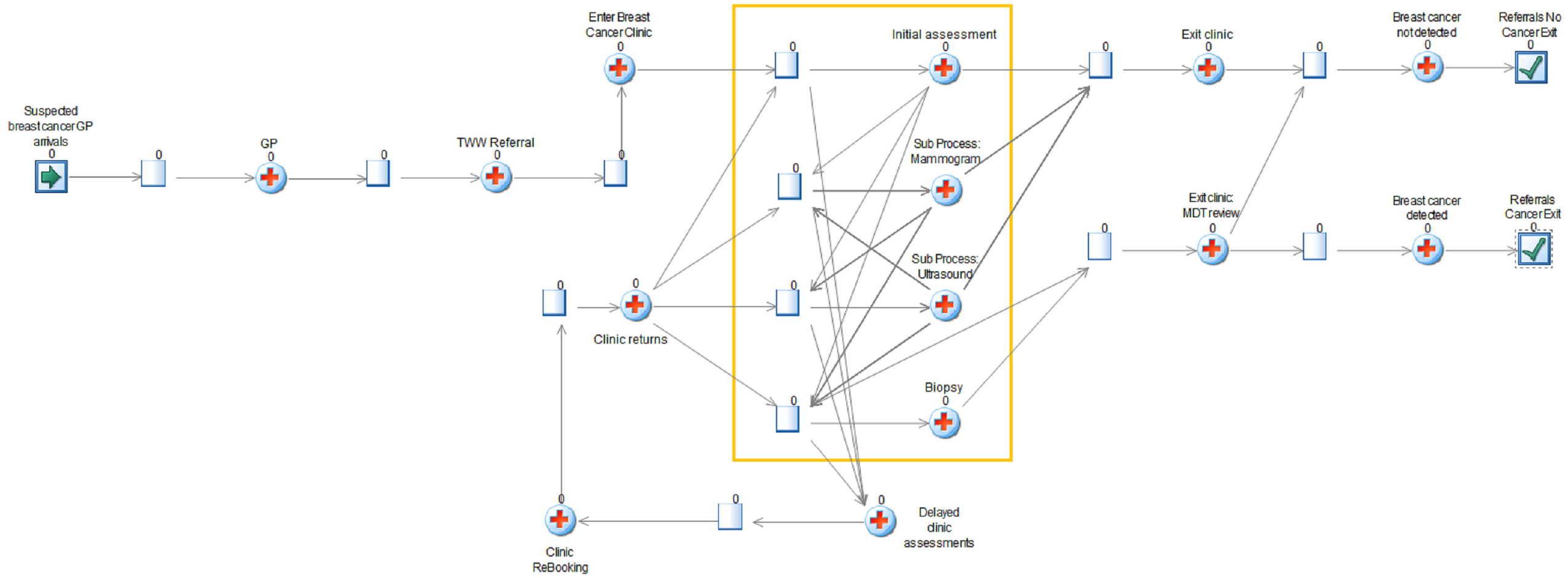
Model figure for the standard care (no primary care testing) pathway (GP = general practitioner; TWW = two week wait; MDT = multi-disciplinary team) Patients with suspected breast cancer enter the model at the GP arrivals point. Individual patients are then tracked through the model. The yellow box captures the activities of the breast cancer clinic (i.e. initial assessment; mammogram; ultrasound and biopsy), which each depend the availability of rooms and staff. Patients are required to wait in the activity queues until such a time as an available room and required staff members are available. At the end of each scheduled clinic, any patients left in the clinic activity queues are routed back around to the ‘Clinic returns’ activity and will be required to attend a future clinic to finish their course of diagnostics. Once an individual’s clinic activities have been completed, the patient receives a diagnosis of cancer or no cancer, and exits the model at the associated exit point.

**Figure 2.**
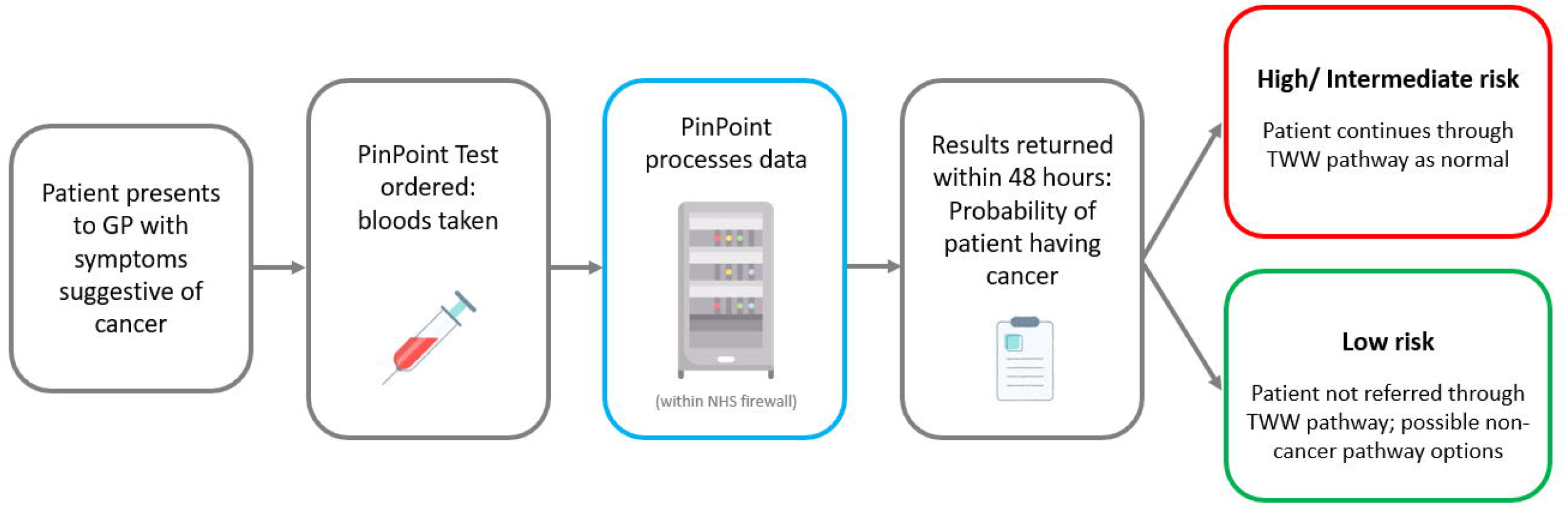
Expected implementation pathway for the PinPoint test (GP = general practitioner; TWW = two week wait).

A warm-up period of twelve weeks is initially applied in the model to populate the service queues (i.e. rather than starting with an empty system). The subsequent results collection period lasts one year, and is intended to capture service demand levels matching those observed at the LTHT clinic in 2019.

### Model Parameters

This section outlines the key model parameters – a full list is provided in **Additional files 1 and 2**. The inter-arrival time of new patients into the model was set to an exponential distribution (mean = 16.2778 minutes), which reflects the annual number of patients seen at the LTHT clinic (n=10,542; based on 2019 clinical audit data provided by the LTHT) assuming a steady arrival of patients. The prevalence of breast cancer was set to 4.76%, based on analysis of a bespoke data extract from the LTHT electronic health record and associated Leeds Patient Level Information and Costing System (PLICS) dataset for 2018/19.

Patients entering the clinic for the first time undergo an initial assessment. All patients with cancer were assumed to be referred for imaging, whilst 25% of patients without cancer were allowed to be discharged at this point (based on the LTHT audit data). The series of diagnostic activities undertaken for imaged patients was set based on the Leeds PLICs dataset: this data showed that, at the LTHT clinic, most patients without breast cancer undergo ultrasound only (39%), or some combination of mammogram and ultrasound (45%); whilst patients diagnosed with cancer most often undergo the full ‘triple assessment’ of mammogram, ultrasound and biopsy (85%) (see **Table 1**). Whilst a minority of patients with cancer were indicated as having *not* undergone biopsy in this dataset (30/362; 8.3%), it was assumed in the model that all patients with cancer *would* require a biopsy (based on clinical expert opinion), and this activity was therefore added to those patients’ sequence of events.

**Table 1.**
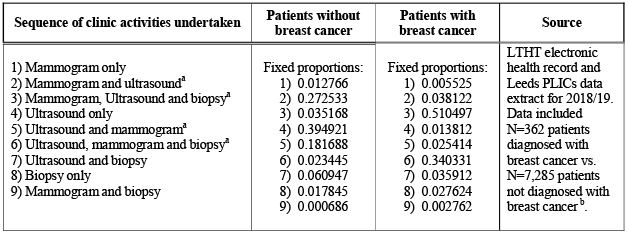
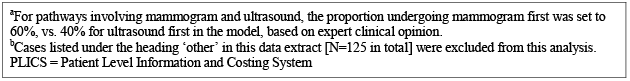
Proportion of patients following different sequences of breast cancer clinic diagnostic activities.

In line with the LTHT clinic schedule, the model includes seven weekly ‘full’ clinics (Mon AM, Tues PM, Weds AM & PM, Thurs AM & PM, and Fri AM), and one weekly ‘add-on’ clinic with a reduced capacity (Tues AM; this clinic was first introduced in early 2018 as a temporary measure to alleviate pressure on the clinic, but has since been permanently adopted due to persisting demand). All clinics last four hours (AM = 09:00 to 13:00; PM = 14:00 to 18:00), with up to fifteen minutes of staff overtime allowed to complete activities if necessary. The clinic staff includes radiological and surgical staff (consultants, nurse practitioners [NPs] and physician associates [PAs]); and radiographer staff (Band 6 sonographers and non-sonographers, Band 7 sonographers and Grade 2 assistants). Staff numbers were set to match the LTHT staff schedule, and staff availability within shifts was set based on expert opinion as to the amount of time each staff member would typically spend on ‘other’ activities (e.g. administrative tasks and follow-up appointments) (see **Additional files 1** and **2**). Patients are not assigned to a specific member of staff – rather, as soon as any relevant staff member becomes available for the patient’s next required activity, the patient can undergo that activity.

Based on consultation with LTHT clinic staff, the default number of new patients booked into each clinic was set to twenty-five for full clinics, and ten for the Tues AM add-on clinic. Whilst these are the ‘target’ clinic numbers, the clinic audit data indicated that the full clinic numbers could fluctuate (little variation was observed in the Tues AM clinic). It was confirmed that full clinic numbers would be increased in response to periods of higher demand, in an attempt to meet the TWW target. In the model therefore, above a clinic queue of 380 (i.e. slightly above the number that can be seen over two weeks at the default clinic numbers), the number of new patients booked into the next full clinic was allowed to increase by 1 for every additional 15 patients in the queue, up to a maximum of 34. The clinic numbers thus reflect a degree of ‘responsiveness’ to the queue, in line with real-life practice. This approach was able to produce similar proportions of patients meeting the TWW target in the model standard care arm (66.3%), as seen in NHS referral data for the Leeds CCG in 2019 (66.5%) (23). All queues in the system were set to operate on a ‘first in first out’ basis – such that those patients waiting the longest in queues are always the first to be selected for the associated activity.

Clinics may close with patients still waiting in the activity queues. In this case, the ‘overspill’ patients are routed out of the clinic and placed in a queue to return to the clinic at a later date. Based on the audit data, five appointments are typically reserved per LTHT full clinic to see overspill patients from previous clinics, with a slightly higher number (six) observed for the Tues AM clinic. The model therefore allows up to five and six overspill patients to be seen per full clinic and Tues AM clinic respectively (in addition to new patients).

The maximum number of each clinic activity able to be undertaken at any given time was based on the number of rooms (each with one available imaging device) available at the LTHT clinic (four initial assessment; three mammogram; four ultrasound; and four biopsies). The actual number of each activity undertaken at any point depends also on the availability of staff: an activity can only go ahead if both a room and required staff are available. The median times taken to complete each activity was based on expert opinion (initial assessment = 10 minutes; mammogram = 20 minutes; ultrasound = 20 minutes; biopsy = 30 or 45 minutes [each with 50% chance of occurring]), allowing for some variation around the expected timings (see **Additional File 1**).

The sensitivity (0.98) and specificity (0.20) values applied for the Pinpoint test were provided by the manufacturer, based on an unpublished test development and validation study. This study included data on a total of n=14,021 patients referred along the TWW pathway for breast cancer at LTHT, between 2011-16 [study development set] and 2017-19 [study validation set]). Full details of this study are now available in a published manuscript (12). Other key parameters for the Pinpoint arm were altered within scenario and secondary analyses, as outlined further below.

### Validation

Face validity of the model was checked via a series of consultations with clinicians at the LTHT clinic. Internal validity of the model code was confirmed using extreme value tests, and using the Simul8 inbuilt ‘Simulation Monitor’ function. Finally, external validity of the model was determined by comparing primary baseline results from the model against real world data: (i) the simulated proportion of patients meeting the TWW target (66.3%) were compared against NHS England data for the same time period (66.5%) (23); and the number of simulated overspill appointments (N=1,685) were compared against the annual value reported in the LTHT audit data (N=1,664).

### Analysis

All results are based on 150 model trials (i.e. running the model 150 times, using different random number sequences), and using deterministic analysis (i.e. probabilistic sensitivity analysis was not conducted due to the early nature of this analysis). The number of trials and warm-up period were set to a level sufficient to produce stable model outputs (ascertained by visual inspection of plotted model outputs).

For Pinpoint, due to current uncertainty as to the exact impact of the test on standard care, two primary scenarios were explored:

- **Scenario 1**: GPs only refer patients with high-risk results (i.e. 100% adherence to testing), and all patients with cancer who receive a low-risk test result are assumed to return after 6 weeks with persisting symptoms and receive a delayed clinic referral.
- **Scenario 2**: GPs overrule 20% of low-risk results (cancerous and non-cancerous cases), and a further 10% of patients without breast cancer with a low-risk test result are also assumed to return and receive a delayed referral at six weeks.

In addition to the above, based on the premise that Pinpoint could significantly reduce referrals and free up clinic capacity, the following service reconfiguration scenarios were explored: (1) removing the Tues AM add-on clinic; (2) removing a full clinic (arbitrarily chosen as Wed AM); and (3) removing the Tues AM and Wed AM clinics. These options were applied together with the Pinpoint Scenario 2 parameters. A range of secondary analyses were also conducted to explore the impact of altering the levels of GP adherence, delayed referrals, diagnostic accuracy, cancer prevalence and patient arrivals (see **Additional File 3**).

## Results

### Referral patterns

Based on average simulated patient numbers over 150 model runs, under standard care, 100% (n=10,528) of patients receive an immediate referral. With Pinpoint, 81% (n=8,544/ 10,550) of patients completing the model were referred under Scenario 1: this included all patients with breast cancer (n=505), 2% (n=10) of whom received a delayed (rather than immediate) referral; and 80% (n=8,039/10,044) of patients without cancer (all referred immediately). Under Scenario 2, 86% (n=9,107/10,550) of patients received a referral: all patients with breast cancer (n=505), 2% (n=8) of whom received a delayed referral; and 86% (n=8,602/ 10,044) of patients without breast cancer, 2% (n=161) of whom also received a delayed referral. Note that in both scenarios, the average number of patients completing the model in the PinPoint arm is higher than in the standard care arm, due to the fact that more patients remain in the system queues (i.e. the TWW referral queue, and the overspill Clinic Returns queue) at the end of the simulation in the standard care arm compared to the PinPoint arm.

### Primary outcomes

**Table 2** shows that the reduction in referrals achieved with the PinPoint test is expected to shorten the time to clinic for those referred, by three to four days on average. Consequently the percentage of patients seen in the clinic in under two weeks is increased, from around 66% with standard care, to between 100 (Scenario 1) and 98% (Scenario 2) with the Pinpoint test. Overspill cases are also reduced – from an average of 1,685 with standard care (a number which closely matches the number of overspill appointments recorded in the LTHT 2019 audit data i.e. n=1,664), to between 727 (Scenario 1) or 886 (Scenario 2) with Pinpoint.

**Table 2.** Model results: primary outcomes for standard care, PinPoint testing scenarios and service reconfiguration scenarios.

| Strategy | Number completing the model<br>(95% CI) |  |  | Average time to clinic<br>[week days]<br>(95% CI) |  | Percentage achieving TWW Target<br>(95% CI) |  |  | Number of<br>Overspill<br>(95% CI) |
| --- | --- | --- | --- | --- | --- | --- | --- | --- | --- |
|  | Total | BC | No BC | BC | No BC | BC | No BC | Total |  |
| **Standard care** | **10,528**<br>(10,511 to 10,545) | **503**<br>(499 to 507) | **10,025**<br>(10,008 to 10,042) | **9.81**<br>(9.81 to 9.82) | **9.81**<br>(9.81 to 9.81) | **66.4%**<br>(66.0 to 66.9) | **66.3%**<br>(66.0 to 66.5) | **66.3%**<br>(66.0 to 66.5) | **1,685**<br>(1,648 to 1,722) |
| **Pinpoint scenarios** |  |  |  |  |  |  |  |  |  |
| **Scenario 1**<br>[0% GP override; 0% true negatives receive delayed referral] | **10,549**<br>(10,532 to 10,567) | **505**<br>(502 to 509) | **10,044**<br>(10,027 to 10,061) | **6.23**<br>(6.20 to 6.25) | **5.64**<br>(5.63 to 5.64) | **98.0%**<br>(97.9 to 98.1) | **100%**<br>(100 to 100) | **99.9%**<br>(99.9 to 99.9) | **727**<br>(707 to 746) |
| **Scenario 2**<br>[20% GP override; 10% true negatives receive delayed referral] | **10,550**<br>(10,532 to 10,567) | **505**<br>(502 to 509) | **10,044**<br>(10,027 to 10,061) | **6.28**<br>(6.25 to 6.31) | **6.36**<br>(6.35 to 6.38) | **98.4%**<br>(98.3 to 98.5) | **98.1%**<br>(98.1 to 98.1) | **98.1%**<br>(98.1 to 98.2) | **886**<br>(861 to 911) |
| **Service reconfiguration scenarios (applied with Pinpoint Scenario 2 parameters)** |  |  |  |  |  |  |  |  |  |
| **No Tues AM clinic**<br>[capacity = 10 new patients + 6 overspill] | **10,497**<br>(10,484 to 10,510) | **502**<br>(499 to 506) | **9,995**<br>(9,982 to 10,008) | **7.75**<br>(7.61 to 7.89) | **7.83**<br>(7.69 to 7.97) | **95.0%**<br>(93.8 to 96.2) | **94.8%**<br>(93.7 to 96.0) | **94.8%**<br>(93.7 to 96.0) | **1,292**<br>(1260 to 1324) |
| **No Weds AM clinic**<br>[capacity = 25 to 34 new patients + 5 overspill] | **10,470**<br>(10,453 to 10,487) | **498**<br>(495 to 502) | **9,972**<br>(9,954 to 9,989) | **11.72**<br>(11.69 to 11.75) | **11.81**<br>(11.79 to 11.82) | **2.9%**<br>(2.3 to 3.4) | **2.9%**<br>(2.3 to 3.4) | **2.9%**<br>(2.3 to 3.4) | **1,632**<br>(1,596 to 1,668) |
| **No Tues AM or Weds AM clinic** | **10,274**<br>(10,275 to 10,291) | **461**<br>(459 to 464) | **9,813**<br>(9,796 to 9,830) | **12.49**<br>(12.46 to 12.52) | **12.58**<br>(12.57 to 12.58) | **0.0%**<br>(0.0 to 0.0) | **0.0%**<br>(0.0 to 0.0) | **0.0%**<br>(0.0 to 0.0) | **2,057**<br>(2,042 to 2,071) |
| BC = breast cancer; CI = confidence interval. |  |  |  |  |  |  |  |  |  |

**Table 2** further illustrates that, applying the Scenario 2 parameters, the reduction in referrals achieved with the Pinpoint test is sufficient to allow the Tues AM add-on clinic to be removed from the weekly clinic schedule: this option maintains the percentage meeting the TWW target at close to 95%, and keeps the overspill cases (n= 1,292) well below that seen with standard care (albeit higher than the PinPoint main scenario analyses). Any further reductions to the schedule are not tolerated however, with the removal of the full Wed AM clinic resulting in less than 3% of patients meeting the TWW requirement and high overspill numbers (n = 1,632).

The results were robust to the secondary analyses explored (see **Additional file 3**). The incremental benefits associated with the PinPoint test were slightly reduced when (a) increasing the proportion of GP overrides up to 50%, or (b) increasing the proportion of non-cancerous patients receiving a delayed referral up to 50%; however the test maintained significantly higher TWW percentage values and lower overspill numbers compared to standard care, across all of the secondary analyses conducted. The test-based strategy was also more robust than standard care at coping with increased patient numbers: for example, when increasing the number of patients arriving in the model by 10%, the PinPoint test was able to maintain 99.9% of patients meeting the TWW target, with an average of 1,156 overspills; whilst the same scenario for standard care led to a drop in the overall percentage meeting the TWW target (to 61.2%), with an average of 2,555 overspills.

## Discussion

Based on early diagnostic accuracy data and clinical assumptions, the PinPoint test could provide significant relief to breast cancer clinics, enabling the majority of referred patients to be seen within two weeks and allowing more patients to receive their full sequence of diagnostic assessments in a single clinic visit. It is further possible that this reduction in clinic demand could be sufficient to remove the LTHT weekly add-on clinic, if the benefits of doing so (e.g. staff relief, cost savings, shifting resources) could be considered to outweigh the associated costs (e.g. slightly lower percentage of patients meeting the TWW target, and higher overspill numbers). It is of particular interest to note that, whilst in simple terms the test is expected to remove 20% of patients from the TWW pathway (based on its 20% specificity), this does not translate to being able to remove 20% of the clinic schedule (i.e. one full clinic). This is because of the significant extra demand in workload the department is already attempting to meet by regularly expanding their clinics to maximum capacity; removing some of that excess demand means that clinics are more able to regularly run at their intended (rather than maximum) capacity.

As with any early model-based analysis, this assessment has limitations. First, the model is based on data from a single centre, and the results are therefore only expected to reflect the impact of the test on similar medium-to-large sized clinics across the UK. The applicability of the findings to other NHS clinics, particularly those of a different size or alternative configurations to the Leeds clinic, is unclear. Second, due to the early nature of this assessment, several of the model parameters were set based on expert opinion. Ideally a site visit to observe the actual flow of patients through the LTHT clinic would have been undertaken, but was not possible due to the ongoing COVID pandemic. Nevertheless, the availability of clinic audit data and the bespoke NHS PLICs dataset for Leeds allowed most parameters to be set or validated against real world data. Most importantly for the PinPoint arm, key assumptions had to be made around the management and behaviour of patients with low-risk results, for which there is currently no available evidence. If and when further information on these parameters becomes available, the model can be easily updated to provide a more precise evaluation.

Additional uncertainty around the applicability of these findings stems from the ongoing fallout of the COVID-19 pandemic, which has had a major impact on cancer referral patterns (24, 25). At the time of analysis planning (mid 2020), the demand for breast cancer clinic services at the LTHT was considered to be unpredictable due to the impact of successive national and regional “lock-downs”. Nevertheless, based on expert consultation, only a slight drop in patient numbers was observed over the initial national lock-down period (early 2020), with numbers expected to quickly return to pre-pandemic levels following continued easing of restrictions. The decision was therefore made to base the analysis on pre-pandemic data from 2019. Going forward, it is expected that referrals will likely rise above pre-pandemic levels. If that expectation holds true, based on our early analysis the PinPoint test would potentially be better enable secondary care breast cancer services to cope with such increases in demand, and could therefore be of increased utility.

Whilst the findings of this exploratory study are encouraging, additional outcomes – most importantly patient health outcomes and health-economic outcomes – would require evaluation before adoption of the PinPoint test could be definitively recommended. Of particular concern is the clinical management and outcomes for those 20% of patients initially ruled-out by the PinPoint test. This group was explored in the current model via sensitivity analyses, which show how the model outcomes may change if a higher proportion of initially discharged patients return to their GP for a later referral.

However, the potential impact on health outcomes for this group were not explored. Although this group consists predominantly of patients without cancer, the significant risks associated with delayed or missed diagnoses for those few patients with cancer is such that some form of safety-netting guidance is expected to be required, in order to instil confidence that patient harm may be avoided. Repeated testing and/or a follow-up GP visit could be implemented in this group, for example, as a means of protection both for those patients with cancer, and for those patients without cancer who may experience persisting symptoms resulting from other indications requiring further follow-up. Clearly the implementation of any such safety-netting procedures would have an impact on both healthcare costs and patient health outcomes. An early economic evaluation is currently underway to explore the impact of different implementation scenarios core to the health-economic argument for the use of the PinPoint test, compared to current practice. The findings of that analysis will help to determine whether further primary research is required in order to address these aspects of uncertainty.

The results of this early-stage evaluation were used to help secure further funding for a service evaluation of the PinPoint test across multiple sites in Yorkshire (26). That project will aim to determine the real-world performance of the test; establish the logistics of implementing the test in the NHS; and update the early evaluation described herein, to address key uncertainties highlighted in the analysis and incorporate cost and health outcomes data. The results of the service evaluation will be reported to NHS England.

On a final note, alternative strategies have been suggested for reducing the burden on secondary care services. Blacker and colleagues (27), for example, suggest that separate non-urgent clinics for breast pain and breast lumps in patients aged under thirty could be set up, based on the low incidence of cancer in this group. Ramzi and colleagues (28) have further demonstrated that using age as a single referral criterion provides higher diagnostic accuracy than the TWW pathway, and would be expected to be less costly. Other multi-variate prediction models, which avoid the need for any additional laboratory-based tests, have also been suggested (29). The use of the PinPoint test as a prioritisation test for referrals (i.e. use-case (2)), in addition to its role as a rule-out test, also deserves further exploration(12). Ideally future studies should consider the comparative utility of alternative rule-out and/or priority-setting referral strategies, so that the most clinically- and cost-effective option may be selected.

## Conclusions

Based on early data, this exploratory analysis found that the PinPoint risk assessment test could help to alleviate pressure on cancer clinics. Further research is now required to assess broader clinical and economic impacts, and to determine outcomes and risks for those patients deemed to be low-risk on the PinPoint test.

## Data Availability

The simulation model generated during the current study is available from the corresponding author upon reasonable request.

## Declarations

### 1. Ethics approval and consent to participate

Not applicable (no human participants were involved in this research).

### 2. Consent for publication

Not applicable.

### 6. Authors contributions

AS conducted the data analysis for the model, developed and parameterised the model, analysed the model results, and was the primary author of the manuscript. SF provided input on the model development and parameterisation. NS provided clinical data for the model, supervised the data analysis, and clinically validated the model. RN provided clinical oversight throughout the development and analysis of the model. LH provided PLICS data for the model and supervised the data analysis. BS provided supervision throughout the model development and analysis. All authors contributed to the drafting and finalisation of the manuscript.

## 7. Acknowledgements

Not applicable.

## List of abbreviations

BC =: breast cancer
CCG =: Clinical Commissioning Group
CI =: confidence interval
DES =: discrete event simulation
LTHT =: Leeds Teaching Hospitals Trust
GP =: General practitioner
MDT =: multi-disciplinary team
NP =: nurse practitioner
PA =: physician associate
PLICS =: Patient Level Information and Costing System
TWW =: two-week wait
UK =: United Kingdom

