## Additional file 1 for "An exploratory assessment of the impact of a novel risk assessment test on breast cancer clinic waiting times and workflow: a discrete event simulation model"

**Additional file 1. Table of PinPoint capacity model parameters**

| **Parameter** | **Value/ Distribution** | **Source/ details** |
| --- | --- | --- |
| **Clock properties** | | |
| Length of day | 11 hours (08:00 to 19:00) | - |
| Length of week | 5 days (Monday to Friday) | - |
| Warm up period | 12 weeks | - |
| Results collection period | One year (52 weeks) | Note: total modelled time = 171,600 minutes [11 hours/day * 5 days/week * 52 weeks]. |
| **Incidence and inter-arrival time** | | |
| Breast cancer incidence | Fixed (0.0476) | LTHT PLICs data for 2018/19 (370/ 7,772 diagnosed with breast cancer). |
| GP inter-arrival time | Exponential (mean = 16.27775) | Based on numbers seen for annual period (Jan-Dec 2019) at LTHT breast cancer clinic (audit data). Total number seen = 10,542. Mean inter-arrival time = total model time (171600) / 10542 = 16.27775 (i.e. one new GP presentation every 16.27775 minutes in the model). |
| **Event durations (minutes)** | | |
| Time in GP | 0 | - |
| Minimum Wait Time in Queue for TWW referral (minutes) | Fixed value (3300) | Assumed a minimum of 1 week (3,300 model minutes) would be required to book patients into the next available clinic. |
| Time in initial assessment | Triangular (8,10,12) | Expert opinion. |
| Time in mammography | Triangular (15,20,25) |  |
| Time in ultrasound | Triangular (15,20,25) |  |
| Time in biopsy | Probability profile: 50% 30; 50% 45 |  |
| Time in patient preparation | Triangular (2,3,4) | Assumption – verified with expert. |
| Minimum Wait Time in Queue for Clinic Returns | Fixed (180) | Expert opinion – returning patients would need to return at least one clinic later (e.g. patients unable to undergo all required activities in an AM clinic would need to wait at least until the following morning to return). Effectively in the model this requires a minimum wait time of 3 hours (180 mins). |
| Time in multidisciplinary team meeting (MDT) review | 0 | - |
| **Clinic capacity and clinical pathways** | | |
| Number of new patients booked in per full clinic (i.e. Mon AM, Tues PM, Weds AM, Weds PM, Thurs AM, Thurs PM, Fri AM) | Dependent on the clinic queue size:  25 if queue ≤ 380;  26 if queue > 380;  27 if queue > 395;  28 if queue > 410;  29 if queue > 425;  30 if queue > 440;  31 if queue > 455;  32 if queue > 470;  33 if queue > 485;  34 if queue > 500. | The default (N=25) and max (N=34) numbers were set based on LTHT clinic 2019 audit data and expert consultation. It was further assumed that: (i) due to the TWW target, the default number would be increased if the queue rose above the total N that could be seen in 2 weeks at the default values (i.e. 25*7 + 10*1 = 370); and (ii) that the number would rise by 1 every +15 patients added to the queue thereafter (determined via consultation with an LTHT clinician). Assigning an additional 1 patient per 7 weekly clinics ensures that an additional 14 patients can be seen over a 2 week period – roughly dealing with the increased demand of 15 extra patients. |
| Maximum number of "overspill" patients seen per full clinic | Fixed value (5) | LTHT breast cancer clinic 2019 audit data and expert consultation. |
| Number of new patients booked in per Tues AM clinic | Fixed value (10) | LTHT breast cancer clinic 2019 audit data and expert consultation. |
| Maximum number of "overspill" patients seen per Tues AM clinic | Fixed value (6) | LTHT breast cancer clinic 2019 audit data and expert consultation. |
| Proportion of patients who undergo initial assessment | Fixed value (100%) | Expert consultation. |
| Proportion of patients without breast cancer referred for imaging | Normal (0.75, 0.06) | LTHT breast cancer clinic 2019 audit data. |
| Proportion of patients with breast cancer referred for imaging | Fixed value (1) | Assumption. |
| No breast cancer cohort: proportion following clinical pathways 1 to 9:  1) Mammogram only  2) Mammogram and ultrasound  3) Mammogram, ultrasound and biopsy  4) Ultrasound only  5) Ultrasound and mammogram  6) Ultrasound, mammogram and biopsy  7) Ultrasound and biopsy  8) Biopsy only  9) Mammogram and biopsy | Fixed values: 1) 0.012766  2) 0.272533  3) 0.035168  4) 0.394921  5) 0.181688  6) 0.023445  7) 0.060947  8) 0.017845  9) 0.000686 | LTHT Leeds PLICs data for 2018/19, combining data on patients referred via the ‘breast cancer symptoms’ pathway and the ‘suspected breast cancer’ pathway.  Data included N=362 patients diagnosed with breast cancer, and N=7,285 patients not diagnosed with breast cancer Cases listed under the heading ‘other’ [N=8 in the breast cancer cohort, and N=117 in the non-cancer cohort] were excluded from this analysis.  For pathways involving mammogram and ultrasound, the proportion undergoing mammogram first was set to 60% vs. 40% for ultrasound first. Current clinical guidelines recommend Mammogram first for women aged 40 and over (1). The proportion of referred women aged 40 and over was estimated as 60% based on expert clinical opinion. |
| No breast cancer cohort proportion following clinical pathways 1 to 9:  1) Mammogram only  2) Mammogram and ultrasound  3) Mammogram, ultrasound and biopsy  4) Ultrasound only  5) Ultrasound and mammograms  6) Ultrasound, mammogram and biopsy  7) Ultrasound and biopsy  8) Biopsy only  9) Mammogram and biopsy | Fixed values:  1) 0.005525  2) 0.038122  3) 0.510497  4) 0.013812  5) 0.025414  6) 0.340331  7) 0.035912  8) 0.027624  9) 0.002762 |  |
| Max number of initial assessments at any one time | Fixed value (4) | Expert opinion. |
| Max number of mammograms at any one time | Fixed value (3) |  |
| Max number of ultrasounds at any one time | Fixed value (4) |  |
| Max number of biopsies at any one time | Fixed value (4) |  |
| **Staff availability and overtime** | | |
| Consultants availability | 83.33% | Expert opinion, based on the amount of time staff are expected to work on other activities per 4-hour clinic:   - Consultants = 40 minutes - NPs & PAs = 1 hour - Band 6 non-sonographers = 30 minutes - Band 6/7 sonographers = 45 minutes - Band 2 assistants = 2 hours   Note: in the model these % availability values are applied to each staff member (i.e. resource). Assigning an availability of, say, 75%, means that that Resource will only be available to work on its designated tasks for 75% of simulation time. |
| Nurse Practitioners (NPs) and Physician Associates (PAs) availability | 75% |  |
| Band 6 non-sonographers availability | 87.5% |  |
| Band 6 and Band 7 sonographers availability | 81.25% |  |
| Band 2 Assistants availability | 50% |  |
| Staff maximum overtime allowance per shift (all staff) | 15 minutes | Assumption, verified with LTHT clinician. Note: each activity in the clinic no longer accepts new patients if the time taken to complete the activity will require staff to stay more than 15 minutes after their shift. |
| **Pinpoint parameters** | | |
| Pinpoint sensitivity | Fixed value (0.98) | Manufacturer data (now available as a pre-print manuscript (2)) |
| Pinpoint specificity | Fixed value (0.20) |  |
| Proportion of low risk results overridden by the GP | Fixed values:  Scenario 1: 0.00  Scenario 2: 0.20 | Modeller assumption. |
| Proportion of patients without breast cancer with a low risk result who return with persisting symptoms | Fixed values:  Scenario 1: 0.00  Scenario 2: 0.10 | Modeller assumption. |
| Proportion of patients with breast cancer with a low risk result who return with persisting symptoms | Fixed value (1.00) | Modeller assumption. |
| Referral delay for patients with persisting symptoms | Fixed value (6 weeks) | Modeller assumption (6 weeks = 19,800 model minutes) |
| LTHT = Leeds Teaching Hospital NHS Trust; PLICS = Patient Level Information and Costing System; TWW = two-week wait; NP = Nurse Practitioner; PA = Physician Associate. | | |
