## Additional file 2 for "An exploratory assessment of the impact of a novel risk assessment test on breast cancer clinic waiting times and workflow: a discrete event simulation model"

**Additional file 2. Table of breast cancer clinic staff numbers applied in the PinPoint capacity model**

| **Clinic** | **Surgical staff** | | | **Radiographer staff** | | |
| --- | --- | --- | --- | --- | --- | --- |
|  | **Consultants** | **NPs** | **PAs** | **Band 6 non-sonographers** | **Band 6 sonographers** | **Band 7 sonographers** |
| MON AM | 3 | 2 | 2 | 5 | 2 | 3 |
| TUES AM | 1 | 0 | 0 | 5 | 2 | 3 |
| TUES PM | 3 | 2 | 1 | 5 | 2 | 3 |
| WEDS AM | 2 | 2 | 2 | 5 | 3 | 5 |
| WEDS PM | 2 | 2 | 2 | 5 | 3 | 5 |
| THURS AM | 2 | 2 | 1 | 6 | 3 | 6 |
| THURS PM | 1 | 2 | 2 | 6 | 3 | 6 |
| FRI AM | 3 | 2 | 1 | 7 | 2 | 5 |
| NP = Nurse Practitioner; PA = Physician Associate. | | | | | | |
