## Additional file 3 for "An exploratory assessment of the impact of a novel risk assessment test on breast cancer clinic waiting times and workflow: a discrete event simulation model"

**Additional file 3. Table of PinPoint capacity model results: primary outcomes for PinPoint testing scenarios and all secondary analyses**

| **Strategy** | **Number completed**  **(95% CI)** | | | **% Referred to clinic (immediate + delayed referrals)** | | **Average time to clinic [week days]**  **(95% CI)** | | **Percentage Achieving TWW Target**  **(95% CI)** | | | **Number of Overspill**  **(95% CI)** |
| --- | --- | --- | --- | --- | --- | --- | --- | --- | --- | --- | --- |
|  | **Total** | **BC** | **No BC** | **BC** | **No BC** | **BC** | **No BC** | **BC** | **No BC** | **Total** |  |
| **Primary analyses:** | | | | | | | | | | | |
| **Standard care** | 10,528  (10,511 to 10,545) | 503  (499 to 507) | 10,025 (10,008 to 10,042) | 100% | 100% | 9.81  (9.81 to 9.82) | 9.81  (9.81 to 9.81) | **66.4%**  (66.0 to 66.9) | **66.3%**  (66.0 to 66.5) | **66.3%**  (66.0 to 66.5) | **1,685**  (1,648 to 1,722) |
| **Pinpoint Scenario 1**  [0% GP override; 0% true negatives receive delayed referral] | 10,549  (10,532 to 10,567) | 505  (502 to 509) | 10,044  (10,027 to 10,061) | 100% | 80% | 6.23  (6.20 to 6.25) | 5.64  (5.63 to 5.64) | **98.0%**  (97.9 to 98.1) | **100%**  (100 to 100) | **99.9%**  (99.9 to 99.9) | **727**  (707 to 746) |
| **Pinpoint Scenario 2** [20% GP override; 10% true negatives receive delayed referral] | 10,550  (10,532 to 10,567) | 505  (502 to 509) | 10,044  (10,027 to 10,061) | 100% | 86% | 6.28  (6.25 to 6.31) | 6.36  (6.35 to 6.38) | **98.4%**  (98.3 to 98.5) | **98.1%**  (98.1 to 98.1) | **98.1%**  (98.1 to 98.2) | **886**  (861 to 911) |
| **Secondary analyses (PinPoint increased GP override scenarios; baseline = 0% GP override):** | | | | | | | | | | | |
| **10% GP override** | 10,550  (10,532 to 10,567) | 505  (502 to 509) | 10,044  (10,027 to 10,061) | 100% | 82% | 6.22  (6.19 to 6.25) | 5.68  (5.67 to 5.68) | **98.2%**  (98.1 to 98.3) | **100%**  (100 to 100) | **99.9%**  (99.9 to 99.9) | **776**  (755 to 797) |
| **20% GP override** | 10,549  (10,532 to 10,567) | 505  (502 to 509) | 10,044  (10,027 to 10,061) | 100% | 84% | 6.21  (6.18 to 6.23) | 5.73  (5.73 to 5.74) | **98.4%**  (98.3 to 98.5) | **100%**  (100 to 100) | **99.9%**  (99.9 to 99.9) | **838**  (817 to 859) |
| **30% GP override** | 10,550  (10,533 to 10,568) | 505  (502 to 509) | 10,045  (10,028 to 10,062) | 100% | 86% | 6.23  (6.20 to 6.26) | 5.82  (5.81 to 5.84) | **98.7%**  (98.6 to 98.8) | **100%**  (100 to 100) | **99.9%**  (99.9 to 99.9) | **886**  (863 to 909) |
| **40% GP override** | 10,546  (10,529 to 10,563) | 505  (502 to 509) | 10,041  (10,024 to 10,058) | 100% | 88% | 6.39  (6.35 to 6.43) | 6.03  (6.00 to 6.07) | **98.8%**  (98.7 to 98.9) | **100%**  (100 to 100) | **99.9%**  (99.9 to 99.9) | **950**  (926 to 974) |
| **50% GP override** | 10,550  (10,533 to 10,568) | 504  (500 to 507) | 10,023  (10,008 to 10,038) | 100% | 90% | 6.95  (6.85 to 7.04) | 6.64  (6.55 to 6.74) | **98.8%**  (98.6 to 99.0) | **99.8%**  (99.6 to 99.9) | **99.7%**  (99.5 to 99.9) | **1,005**  (982 to 1,029) |
| **Secondary analyses (increasing true negative cases returning with persisting symptoms and a receiving delayed referral; baseline = 0%):** | | | | | | | | | | | |
| **10% true negatives receive delayed referral** | 10,550  (10,533 to 10,568) | 505  (502 to 509) | 10,045  (10,028 to 10,062) | 100% | 82% | 6.26  (6.24 to 6.29) | 6.42  (6.41 to 6.43) | **98.0%**  (97.9 to 98.1) | **97.5%**  (97.5 to 97.6) | **97.6%**  (97.5 to 97.6) | **766**  (748 to 784) |
| **20% true negatives receive delayed referral** | 10,550  (10,533 to 10,567) | 505  (502 to 509) | 10,045  (10,028 to 10,062) | 100% | 84% | 6.32  (6.29 to 6.35) | 7.17  (7.15 to 7.18) | **98.0%**  (97.9 to 98.1) | **95.2%**  (95.2 to 95.3) | **95.4%**  (95.3 to 95.4) | **844**  (823 to 864) |
| **30% true negatives receive delayed referral** | 10,551  (10,534 to 10,568) | 505  (502 to 509) | 10,046  (10,029 to 10,063) | 100% | 86% | 6.41  (6.38 to 6.45) | 7.93  (7.91 to 7.94) | **98.0%**  (97.9 to 98.1) | **93.0%**  (93.0 to 93.0) | **93.3%**  (93.2 to 93.3) | **897**  (872 to 922) |
| **40% true negatives receive delayed referral** | 10,546  (10,529 to 10,563) | 505  (501 to 509) | 10,041  (10,025 to 10,058) | 100% | 88% | 6.61  (6.57 to 6.66) | 8.76  (8.72 to 8.79) | **98.0%**  (98.0 to 98.1) | **90.9%**  (90.8 to 90.9) | **91.3%**  (91.2 to 91.3) | **962**  (938 to 985) |
| **50% true negatives receive delayed referral** | 10,520  (10,506 to 10,535) | 504  (500 to 507) | 10,017  (10,002 to 10,031) | 100% | 90% | 7.20  (7.10 to 7.29) | 9.94  (9.85 to 10.03) | **97.8%**  (97.7 to 98.0) | **88.7%**  (88.5 to 88.8) | **89.2%**  (89.0 to 89.3) | **1,009**  (984 to 1,034) |
| **Secondary analyses (PinPoint reduced clinical performance; default sensitivity = 0.98, default specificity = 0.20):** | | | | | | | | | | | |
| **Sensitivity = 0.931**  **(5% reduction)** | 10,550  (10,533 to 10,567) | 506  (502 to 509) | 10,044  (10,027 to 10,061) | 100% | 80% | 7.68  (7.63 to 7.73) | 5.64  (5.63 to 5.64) | **93.2%**  (93.0 to 93.3) | **100%**  (100 to 100) | **99.6%**  (99.6 to 99.6) | **735**  (715 to 755) |
| **Sensitivity = 0.882**  **(10% reduction)** | 10,550  (10,533 to 10,568) | 506  (503 to 510) | 10,044  (10,027 to 10,061) | 100% | 80% | 9.11  (9.04 to 9.18) | 5.64  (5.63 to 5.64) | **88.4%**  (88.2 to 88.7) | **100%**  (100 to 100) | **99.3%**  (99.3 to 99.3) | **738**  (719 to 757) |
| **Sensitivity = 0.833**  **(15% reduction)** | 10,550  (10,532 to 10,567) | 506  (503 to 510) | 10,044  (10,027 to 10,061) | 100% | 80% | 10.57  (10.49 to 10.64) | 5.64  (5.63 to 5.64) | **83.6%**  (83.3 to 83.8) | **100%**  (100 to 100) | **99.0%**  (99.0 to 99.0) | **742**  (723 to 761) |
| **Specificity = 0.19**  **(5% reduction)** | 10,550  (10,532 to 10,567) | 505  (502 to 509) | 10,044  (10,027 to 10,062) | 100% | 81% | 6.24  (6.22 to 6.27) | 5.66  (5.65 to 5.66) | **98.0%**  (97.9 to 98.1) | **100%**  (100 to 100) | **99.9%**  (99.9 to 99.9) | **766**  (746 to 783) |
| **Specificity = 0.18**  **(10% reduction)** | 10,550  (10,533 to 10,567) | 505  (502 to 509) | 10,045  (10,027 to 10,062) | 100% | 82% | 6.26  (6.23 to 6.29) | 5.68  (5.67 to 5.68) | **98.0%**  (97.9 to 98.1) | **100%**  (100 to 100) | **99.9%**  (99.9 to 99.9) | **773**  (754 to 792) |
| **Specificity = 0.17**  **(15% reduction)** | 10,550  (10,532 to 10,567) | 505  (502 to 509) | 10,044  (10,027 to 10,062) | 100% | 83% | 6.25  (6.23 to 6.28) | 5.65  (5.64 to 5.65) | **98.0%**  (97.9 to 98.0) | **100%**  (100 to 100) | **99.8%**  (99.8 to 99.8) | **797**  (777 to 817) |
| **Secondary analyses (alternative prevalence scenarios; default prevalence = 4.76%):** | | | | | | | | | | | |
| **Standard care arm: prevalence = 3%** | 10,534  (10,517 to 10,551) | 319  (316 to 322) | 10,216  (10,199 to 10,233) | 100% | 100% | 9.81  (9.81 to 9.82) | 9.81  (9.81 to 9.81) | **66.1%**  (65.6 to 66.6) | **66.3%**  (66.0 to 66.5) | **66.3%**  (66.0 to 66.5) | **1,000**  (974 to 1,025) |
| **PinPoint arm: prevalence = 3%** | 10,549  (10,532 to 10,567) | 320  (317 to 323) | 10,229  (10,212 to 10,246) | 100% | 80% | 6.22  (6.18 to 6.25) | 5.63  (5.63 to 5.64) | **98.0%**  (97.9 to 98.2) | **100%**  (100 to 100) | **99.9%**  (99.9 to 99.9) | **391**  (380 to 401) |
| **Standard care arm: prevalence = 7%** | 10,550  (10,533 to 10,568) | 708  (705 to 712) | 9,668 (9,653 to 9,684) | 100% | 100% | 9.81  (9.81 to 9.82) | 9.81  (9.81 to 9.82) | **66.3%**  (66.0 to 66.6) | **66.2%**  (66.0 to 66.4) | **66.2%**  (66.0 to 66.4) | **2,604**  (2,584 to 2,623) |
| **PinPoint arm: prevalence = 7%** | 10,550  (10,533 to 10,568) | 742  (737 to 746) | 9,808  (9,792 to 9,825) | 100% | 80% | 6.25  (6.23 to 6.28) | 5.65  (5.64 to 5.65) | **98.0%**  (97.9 to 98.0) | **100%**  (100 to 100) | **99.8%**  (99.8 to 99.8) | **1,476**  (1,444 to 1,507) |
| **Secondary analyses (increasing numbers arriving; default mean interarrival time [λ] = 16.27775):** | | | | | | | | | | | |
| **Standard care arm: λ = 15.50262** [5% increase in annual number arriving] | 11,025  (11,009 to 11,041) | 524  (520 to 527) | 10,501  (10,485 to 10,518) | 100% | 100% | 9.85  (9.85 to 9.85) | 9.85  (9.85 to 9.85) | **63.4%**  (63.0 to 63.7) | **63.4%**  (63.2 to 63.6) | **63.4%**  (63.2 to 63.6) | **2,238**  (2,209 to 2,267) |
| **PINPOINT arm: λ = 15.50262** [5% increase in annual number arriving] | 11,079  (11,061 to 11,097) | 531  (527 to 534) | 10,549  (10,531 to 10,566) | 100% | 80% | 6.33  (6.30 to 6.36) | 5.74  (5.73 to 5.75) | **98.0%**  (98.0 to 98.1) | **100%**  (100 to 100) | **99.9%**  (99.9 to 99.9) | **925**  (902 to 948) |
| **Standard care arm: λ = 14.79795** [10% increase in annual number arriving] | 11,400  (11,384 to 11,416) | 528  (525 to 531) | 10,872  (10,855 to 10,889) | 100% | 100% | 9.88  (9.88 to 9.85) | 9.88  (9.88 to 9.85) | **61.1%**  (60.7 to 61.4) | **61.2%**  (61.0 to 61.3) | **61.2%**  (61.0 to 61.3) | **2,555**  (2,538 to 2,573) |
| **PINPOINT arm: λ = 14.79795** [10% increase in annual number arriving] | 11,599  (11,582 to 11,617) | 555  (552 to 559) | 11,044  (11,027 to 11,384) | 100% | 80% | 6.71  (6.66 to 6.76) | 6.13  (6.09 to 6.17) | **98.1%**  (98.0 to 98.2) | **100%**  (100 to 100) | **99.9%**  (99.9 to 99.9) | **1,156**  (1,127 to 1,185) |
| **Standard care arm: λ = 14.15456** [15% increase in annual number arriving] | 11,726  (11,708 to 11,745) | 524  (521 to 527) | 11,202  (10,183 to 10,221) | 100% | 100% | 9.92  (9.92 to 9.93) | 9.92  (9.92 to 9.92) | **57.6%**  (57.1 to 58.0) | **57.8%**  (57.5 to 58.1) | **57.8%**  (57.5 to 58.1) | **2,773**  (2,756 to 2,790) |
| **PINPOINT arm: λ = 14.15456** [15% increase in annual number arriving] | 12,045  (12,028 to 12,064) | 576  (572 to 579) | 11,469  (11,452 to 11,486) | 100% | 80% | 9.29  (9.17 to 9.41) | 8.71  (8.59 to 8.83) | **87.4%**  (86.3 to 88.4) | **89.0%**  (88.0 to 90.0) | **88.9%**  (87.9 to 89.9) | **1,398**  (1,364 to 1,431) |
| GP = general practitioner; BC = breast cancer; 95% CI = ninety-five percent confidence interval.  Note: unless otherwise specified, all PinPoint secondary analyses were conducted using the Scenario 1 parameters (i.e. 0% GP override, and 0% true negatives return for a delayed referral) | | | | | | | | | | | |
