## Additional file 4 for "An exploratory assessment of the impact of a novel risk assessment test on breast cancer clinic waiting times and workflow: a discrete event simulation model"

**Strengthening the Reporting of Empirical Simulation Studies (STRESS)**

**Discrete-event simulation guidelines STRESS-DES**

| **Section/Subsection** | **Item** | **Details/ location in Manuscript where details can be found** | |
| --- | --- | --- | --- |
| 1. **Objectives** |  |  | |
| Purpose of the model | 1.1 | The purpose of the model is described in the manuscript Background section. | |
| Model Outputs | 1.2 | The primary model outputs are described in the Background and Methods sections. | |
| Experimentation Aims | 1.3 | The experimental aims of the model are described in the Methods section. In particular, due to the early nature of the evaluation and current uncertainty around the PinPoint test, two scenarios for the PinPoint strategy were considered in the experimental analysis (in addition to sensitivity analyses) – these are detailed in the Method sections. | |
| 2. **Logic** |  |  | |
| Base model overview diagram | 2.1 | The model structure is illustrated in Figures 1 and 2, together with a textual description of the standard care and PinPoint model pathways in the Methods section. | |
| Base model logic | 2.2 | Details of the model logic are provided in the Methods section. | |
| Scenario logic | 2.3 | The scenarios explored are described in the Methods section. | |
| Algorithms | 2.4 | The process underlying the booking-in of patients to the clinic is described briefly in the Methods section, and in more detail in Additional File 1 (under ‘Clinic capacity and clinical pathways’) | |
| Components | 2.5 | 2.5.1 Entities | The model entities (patients) are described in the Methods section. The key attribute applied to patients in the model is breast cancer status, which is defined according to the breast cancer incidence rate provided in Additional File 1. |
|  |  | 2.5.2 Activities | The model activities are described in the Methods section, and illustrated in Figures 1 and 2. Logic underlying when and how patients can enter into activities is described in the Methods section and in Additional File 1. |
|  |  | 2.5.3 Resources | Resources (i.e. staff and rooms) are detailed in the Methods section and further information provided in Additional Files 1 and 2. |
|  |  | 2.5.4 Queues | The queuing discipline (firs in first out) is described in the Methods section. |
|  |  | 2.5.5 Entry/Exit Points | Arrival and exit points of the model are illustrated in Figure 1. |
| 3. **Data** |  |  | |
| Data sources | 3.1 | Data sources are provided in Additional File 1. | |
| Pre-processing | 3.2 | Analysis was undertaken on the LTHT clinic audit data – this consisted of simple tallying and averaging of clinic numbers, overspill appointments, and immediate discharges, to derive key parameters as listed in Additional File 1. | |
| Input parameters | 3.3 | Input parameters are summarised in the Methods section, and provided in Additional File 1. | |
| Assumptions | 3.4 | Any assumptions applied in the model have been clearly highlighted in the Methods section and in Additional File 1. | |
| 4. **Experimentation** |  |  | |
| Initialisation | 4.1 | The warm up period and length of analysis are reported in the Methods section and in Additional File 1. | |
| Run length | 4.2 | The run length of the simulation is provided in the Methods section and Additional File 1. | |
| Estimation approach | 4.3 | The number of replications (trials) and justification for this is reported in the Methods section. | |
| 5. **Implementation** |  |  | |
| Software or programming language | 5.1 | The model was constructed in Simul8, as reported in the Methods section. | |
| Random sampling | 5.2 | Random sampling in the model uses Simul8’s default ‘trials’ random number sequencing. | |
| Model execution | 5.3 | All queues in the model work on a first in first out basis (as reported in the Methods section), such that those patients who have been waiting the longest will always be selected first for activities when resources become available. | |
| System Specification | 5.4 | The model was run on a Lenevo Thinkpad Laptop X280 (intel Core i5, 8^th^ generation), and took approximately 20-30 minutes to run (depending on the scenario evaluated). | |
| 6. **Code Access** |  |  | |
| Computer Model Sharing Statement | 6.1 | Simul8 software can be purchased via the Simul8 website: [https://www.simul8.com](https://www.simul8.comD). Model code can be requested via the corresponding author at:. | |
